# Daily Exceedance of World Health Organization Air Pollutant Guidelines and Cognitive Aging

**DOI:** 10.64898/2026.09.23.26363787

**Authors:** Mikaela Bloomberg, Giorgio Di Gessa, Shaun Scholes, Antonio Gasparrini, Arturo de la Cruz Libardi, Andrew Steptoe, Paola Zaninotto

## Abstract

**Importance:** Long-term exposure to higher average air pollutant concentrations is associated with poorer cognitive health, but annual averages can obscure substantial differences in day-to-day exposure. Whether recurrent high pollution days are associated with long-term cognitive aging trajectories among people with similar overall air pollution exposure is unclear.

**Objective:** To determine whether the annual number of days exceeding World Health Organization daily guideline values for nitrogen dioxide (NO_2_) and particulate matter (PM_2.5_ and PM_10_) is associated with subsequent cognitive decline after accounting for annual mean air pollutant exposure.

**Design:** Prospective cohort study using data from waves 1-10 (2002-03 to 2021-23) of the English Longitudinal Study of Ageing.

**Setting:** Population-based cohort of community-dwelling adults in England.

**Participants:** 12,235 adults aged 50+ years at analytic baseline.

**Exposures:** Number of days exceeding World Health Organization daily guideline values (‘exceedance days’) for NO_2_, PM_2.5_, and PM_10_, derived from residence-linked environmental estimates during the calendar year preceding the analytic baseline.

**Main Outcomes and Measures:** Baseline cognitive performance and model-estimated 14-year cognitive decline (corresponding to the 75th percentile of follow-up duration) in episodic memory and verbal fluency.

**Results:** Exceedance days were not associated with baseline memory or verbal fluency after adjustment for annual mean air pollutant exposure. More NO_2_ exceedance days were associated with faster cognitive decline: comparing the 75th percentile with the median of NO_2_ exceedance days, the difference in 14-year decline was -0.09 SD (95% CI, -0.17 to -0.01; p = 0.02) for memory and -0.10 SD (95% CI, -0.19 to 0.00; p = 0.04) for fluency. PM_2.5_ and PM_10_ exceedance days were not associated with cognitive decline.

**Conclusions and Relevance:** In this cohort study of older adults, a higher number of NO_2_ exceedance days was associated with faster subsequent cognitive decline after adjustment for annual mean air pollutant exposure. This suggests that daily exposure to high NO_2_ concentrations may be related to accelerated cognitive aging even when offset by lower exposure on other days, potentially providing a new rationale for identifying and prioritizing areas with recurrent high NO_2_ exposure and the local emission sources contributing to it.

**Key points:** *Question:* Among individuals with similar average air pollution exposure but different day-to-day exposure patterns, are more high pollution days associated with faster cognitive decline?

*Findings:* In this UK-based cohort of 12,235 adults aged 50+, individuals exposed to more days on which nitrogen dioxide (NO_2_) concentrations exceeded World Health Organization guideline values subsequently had significantly faster cognitive decline over up to 18 years, after adjustment for annual mean air pollutant exposure. No associations were observed for particulate matter.

*Meaning:* Daily exposure to high NO_2_ concentrations is associated with faster cognitive decline even when offset by lower concentrations on other days.

## Introduction

Dementia is a leading contributor to disability, dependency, and health care demand, with an estimated 56.9 million prevalent cases globally.^1^ Because effective treatment options remain limited,^2^ prevention is central to reducing this burden, making the identification of modifiable risk factors for later-life cognitive decline essential. Air pollution is one important late-life risk factor for cognitive health across global settings.^3^ Among the most studied pollutants are nitrogen dioxide (NO_2_), a gaseous pollutant commonly produced during fossil fuel combustion, and particulate matter (PM), a mixture of solid and liquid particles from multiple natural and anthropogenic sources, commonly categorized by aerodynamic diameter into PM_2.5_ (diameter ≤2.5 µm) and PM_10_ (diameter ≤10 µm). Long-term exposure to higher concentrations of NO_2_ and PM_2.5_ has consistently been associated with cognitive decline and dementia risk; evidence for PM_10_ is more limited but also suggests potential adverse associations with cognitive function.^4^

Cognitive health research has primarily focused on average exposure to common air pollutants over months or years. However, this focus may obscure differences in how exposure is distributed over time. Two populations with similar annual mean pollution exposure can experience distinctly different daily exposure patterns: one might experience relatively consistent daily pollution levels, whereas the other could have repeated days of high pollution offset by days of lower pollution concentrations. These exposure patterns may have different implications for cognitive health, as previous studies suggest that short-term peaks in air pollution over days or hours are followed by worse cognitive performance in middle-aged and younger adults.^5–7^ However, whether differences in daily exposure patterns are associated with longer-term cognitive aging has not yet been explored. If repeated high pollution days contribute to cognitive decline among people with similar annual mean exposure, this would provide new support for policies that identify and prioritize local pollution hotspots where recurrent high exposure episodes persist.

In the present study, we used data from 12,235 participants aged ≥50 years in the English Longitudinal Study of Ageing (ELSA) to examine whether daily exposure patterning was associated with cognitive aging over up to 18 years. For each participant, we calculated the number of days exceeding WHO daily guideline values for NO_2_, PM_2.5_, and PM_10_ in the calendar year before analytic baseline, then estimated associations with subsequent cognitive trajectories after adjustment for annual mean exposure to all three air pollutants.

## Methods

### Data sources

ELSA^8^ is an ongoing nationally representative cohort study of community-dwelling adults aged ≥50 years and their partners living in England, with biennial data collection since 2002 and regular refreshment cohorts to maintain national representativeness. Details are published elsewhere.^9^ ELSA received ethics approval from the South Central – Berkshire Research Ethics Committee. Informed consent was obtained at each interview.

ELSA includes cognitive assessments at all waves, with residence-linked environmental exposure estimates available from 2003-2021. The present study used data from waves 1-10 (2002-03 to 2021-23) from core and refreshment ELSA cohorts, the period over which cognitive assessments overlapped with the environmental linkage. Environmental exposure data were available for calendar years corresponding to participants’ interview waves.

Participants were eligible if they had at least one cognitive assessment and had also participated in the immediately preceding ELSA wave. For each participant, the first cognitive assessment meeting this criterion was defined as the analytic baseline, and the immediately preceding wave was defined as pre-baseline. The primary air pollution exposure was calculated once, for the calendar year immediately before the analytic baseline wave and was then related to cognitive trajectories from analytic baseline onward. Covariates were drawn from the pre-baseline wave. This design ensured that covariates preceded exposure, exposure measurement preceded cognitive follow-up, and the timing of exposure assessment relative to analytic baseline was comparable across participants (eFigure 1).

**Figure 1.**
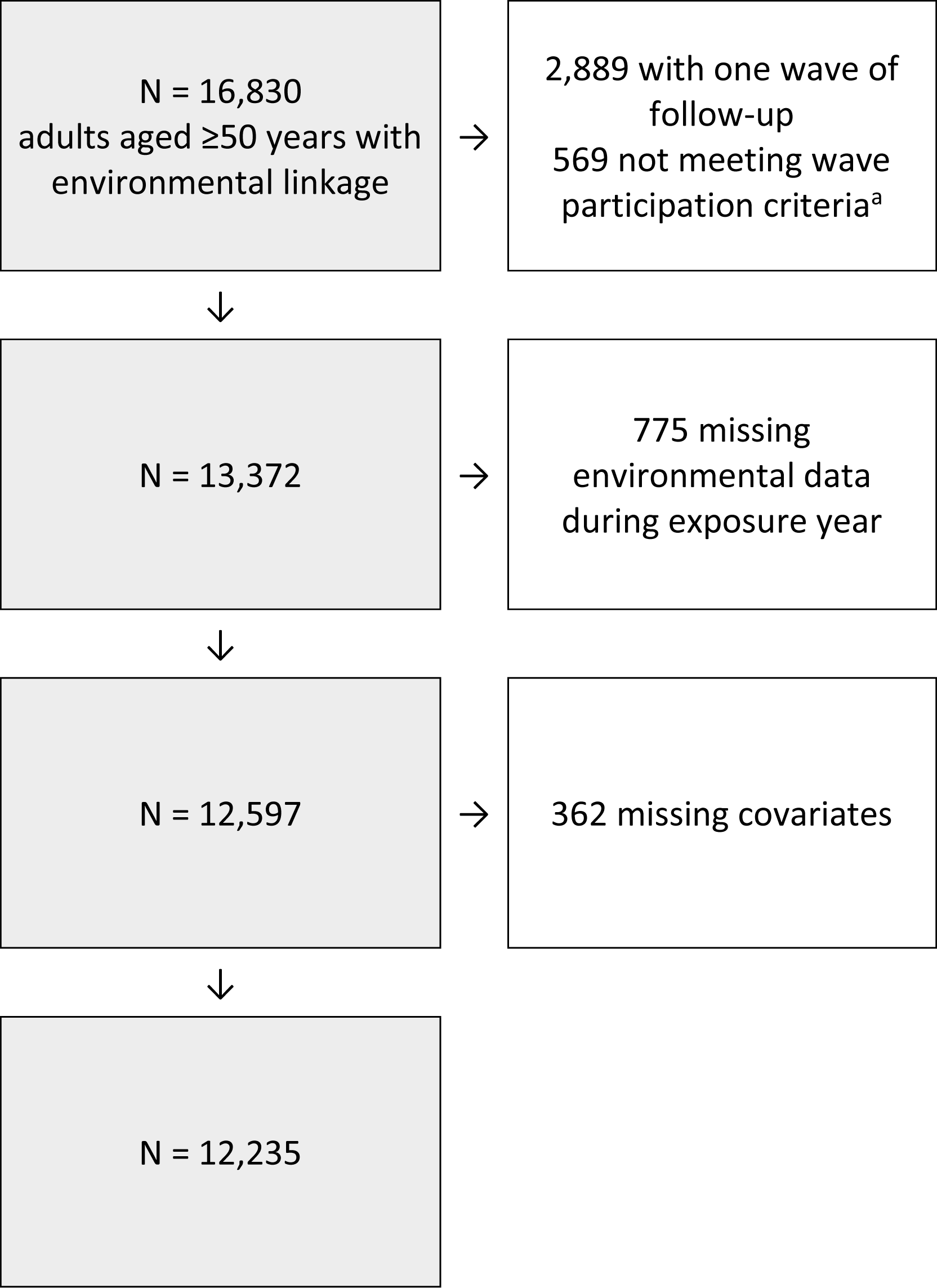
Flowchart of sample selection. ^a^Participants had to have at least one wave of cognitive assessment plus participation in the immediately preceding ELSA wave.

### Air pollutants

The procedure for estimating air pollutants has been described in detail elsewhere.^10^ In brief, daily estimates for NO_2_, PM_2.5_, and PM_10_ were generated on a 1 km x 1 km grid across Great Britain from 2003 to 2021 using a hybrid spatiotemporal machine learning framework, which combines multiple algorithms trained on monitoring station data and spatial and temporal predictors such as emission and dispersion modeling and land use characteristics.^11^ Grid-level estimates were assigned to participants’ residential locations using bilinear interpolation from the four nearest grid cells.

Using these residence-linked daily estimates, we derived exposure measures for the calendar year before analytic baseline. For each participant, we calculated the number of days exceeding WHO daily guideline values^12^ (25 µg/m^3^ for NO_2_, 15 µg/m^3^ for PM_2.5_, and 45 µg/m^3^ for PM_10_) for at least one of the three pollutants, and for each pollutant separately (referred to hereafter as ‘exceedance days’). We also calculated participant-specific annual mean concentrations for each pollutant during the same calendar year. The selected year was intended to characterize participants’ broader pre-follow-up exposure pattern rather than a discrete causal exposure window.

### Cognitive assessment

The cognitive domains examined were episodic memory (assessed using immediate and delayed recall tasks^14^) and verbal fluency (assessed using the animal naming task^15^), which show decline with dementia and are important for daily function.^13^ Details are available in eMethods 1. Cognitive scores were standardized using the distribution at the analytic baseline.

### Covariates

Covariates were selected based on associations with ambient air pollution and cognitive function and were drawn from the pre-baseline wave for each participant. Demographic variables included age in years, birth year, and sex (male or female). Socioeconomic variables included the index of multiple deprivation^16^ (IMD; eMethods 2) to measure neighborhood deprivation, which was categorized into quintiles from least to most deprived, education level (less than secondary, secondary, or above secondary), and net non-pension wealth.^17^ Wealth was standardized by year and categorized into quintiles, with the first quintile corresponding to the least wealth. Finally, to account for spatial confounding, we included urbanicity (urban, town and fringe, village/hamlet/isolated dwelling). Participants with missing pre-baseline covariate data were excluded from the analysis as covariate missingness was minor (<3.0%).

### Statistical methods

We used linear mixed models to examine associations of exceedance days with cognitive trajectories. These models use all available observations and accommodate missingness in the longitudinal outcome assuming a missing-at-random mechanism.^18^ Time was modelled as years since the analytic baseline. Models included random intercepts and slopes for time at the lower-tier local authority (LTLA) level and at the participant level. LTLAs are local government administrative areas were included to account for spatial clustering and residual area-level confounding; the analytic sample included 309 LTLAs. Participants were nested within LTLAs, with unstructured covariance matrices at both levels.

First, we determined the functional form of memory and fluency decline over time using preliminary models that included linear, quadratic, and cubic time terms and pre-baseline age only. Based on visual inspection of fitted trajectories and Wald tests of higher-order time terms, linear and quadratic time terms were retained in memory models and linear, quadratic, and cubic time terms were retained in fluency models. These models were also used to estimate average memory and fluency decline in the analytic sample to provide context for interpreting the magnitude of differences in decline associated with exceedance days.

We then fitted fully adjusted models, with separate models for each exposure (exceedance days in at least one pollutant, NO_2_, PM_2.5_, or PM_10_) and each cognitive domain. These models included exceedance days (modelled as a continuous variable) and interactions between exceedance days and time, capturing associations with cognitive performance and cognitive decline. Fully adjusted models also included participant-specific annual mean NO_2_, PM_2.5_ and PM_10_ concentrations for the exposure year, pre-baseline age, birth year, sex, education, wealth quintile, IMD quintile, urbanicity, and interactions of these variables with time. Associations with exceedance days were therefore estimated conditional on annual mean exposure to all three pollutants. Full model specifications and the model specification process are detailed in eTable 1 and eMethods 3, respectively.

To facilitate interpretation, we estimated model-predicted differences in cognitive performance at analytic baseline and in cognitive decline over 14 years (the 75^th^ percentile of follow-up duration) between the 75^th^ percentile and median of exceedance days for each pollutant and cognitive test, representing a commonly observed contrast in the study population. We also plotted predicted cognitive trajectories at these values, averaged over the observed covariate distribution. Analyses were performed using StataNow 19 MP. Two-sided p-values <0.05 were considered statistically significant.

### Additional analyses

First, we assessed whether results differed by sex or baseline cognitive status. Then, we repeated analyses adjusted for self-reported physician-diagnosed chronic conditions and health behaviors; these variables were not included in the main models because they are unlikely to determine ambient air pollution at participants’ residential locations and are therefore not clear confounders of the associations of interest. Finally, we repeated analyses adjusting for time-varying air pollutant concentrations during the cognitive follow-up period to determine whether post-baseline exposure to air pollutants accounted for the results. Details are provided in eMethods 4.

## Results

### Sample selection and participant characteristics

The final analytic sample included 12,235 participants; sample selection is shown in Figure 1. The mean pre-baseline age in the analytic sample was 62.5 years (SD, 9.6). Of the 12,235 participants, 5,541 (45.3%) were male and 6,694 (54.7%) were female. The median (IQR) duration of cognitive follow-up was 8.3 years (3.7-13.8), with a maximum follow-up period of 18.7 years. Participants who were excluded from the analysis were broadly similar to those included in the analytic sample, though somewhat more socioeconomically disadvantaged (eTable 2).

The mean (SD) annual air pollutant concentration during the exposure year was 25.2 µg/m^3^ (8.2) for NO_2_, 11.7 µg/m^3^ (1.6) for PM_2.5_, and 22.6 µg/m^3^ (3.2) for PM_10_; correlation across the three pollutants is presented in eTable 3. The median (IQR) for exceedance days during the exposure year was 170 days (121-222) for any pollutant, 149 days (90-212) for NO_2_, 86 days (68-94) for PM_2.5_, and 21 days (8-27) for PM_10_. The mean participant-specific SD of daily concentrations during the exposure year was 11.0 µg/m^3^ for NO_2_, 5.6 µg/m^3^ for PM_2.5_, and 9.9 µg/m^3^ for PM_10_ Pre-baseline participant characteristics are presented in Table 1, stratified by percentiles (<25^th^ percentile; 25^th^-75^th^ percentile; >75^th^ percentile) of exceedance days for any of the three air pollutants. Median cognitive follow-up periods were generally similar across levels of exceedance days (8.1 years, IQR 4.0-12.0 for <25^th^ percentile; 9.4 years, IQR 3.7-14.0 for 25^th^-75^th^ percentile; 8.3 years, IQR 2.5-14.0 for >75^th^ percentile).

**Table 1.** Pre-baseline participant characteristics.

|  | Exceedance days (any pollutant) |  |  | P-value |
| --- | --- | --- | --- | --- |
|  | <25<sup>th</sup> percentile | 25<sup>th</sup>-75<sup>th</sup> percentile | >75<sup>th</sup> percentile |  |
| Age in years, mean (SD) | 60.6 (9.0) | 63.1 (9.6) | 63.3 (9.9) | <0.001 |
| Birth year, median (IQR) | 1947 (1938-54) | 1941 (1932-48) | 1941 (1932-48) | <0.001 |
| Sex |  |  |  |  |
| Male | 1373 (45.1) | 2815 (45.7) | 1353 (44.7) | 0.64 |
| Female | 1670 (54.9) | 3348 (54.3) | 1676 (55.3) |  |
| Education level |  |  |  |  |
| Less than secondary | 1032 (33.9) | 2637 (42.8) | 1410 (46.6) | <0.001 |
| Secondary | 1438 (47.3) | 2702 (43.8) | 1168 (38.6) |  |
| Above secondary | 573 (18.8) | 824 (13.4) | 451 (14.9) |  |
| Wealth quintile |  |  |  |  |
| 1 (least wealth) | 464 (15.2) | 1186 (19.2) | 727 (24.0) | <0.001 |
| 2 | 666 (21.9) | 1374 (22.3) | 609 (20.1) |  |
| 3 | 643 (21.1) | 1230 (20.0) | 525 (17.3) |  |
| 4 | 652 (21.4) | 1238 (20.1) | 549 (18.1) |  |
| 5 (most wealth) | 618 (20.3) | 1135 (18.4) | 619 (20.4) |  |
| Neighborhood deprivation |  |  |  |  |
| 1 (least deprived) | 504 (16.6) | 1453 (23.6) | 425 (14.0) | <0.001 |
| 2 | 617 (20.3) | 1277 (20.7) | 428 (14.1) |  |
| 3 | 792 (26.0) | 1123 (18.2) | 451 (14.9) |  |
| 4 | 713 (23.4) | 1118 (18.1) | 671 (22.2) |  |
| 5 (most deprived) | 417 (13.7) | 1192 (19.3) | 1054 (34.8) |  |
| Urbanicity |  |  |  |  |
| Urban | 1371 (45.1) | 4782 (77.6) | 2965 (97.9) | <0.001 |
| Town and fringe | 653 (21.5) | 766 (12.4) | 31 (1.0) |  |
| Village/hamlet/isolated dwelling | 1019 (33.5) | 615 (10.0) | 33 (1.1) |  |

N (%) shown unless otherwise indicated. Exceedance days refer to days during the exposure year exceeding WHO guideline values for at least one of NO<sub>2</sub> (25 µg/m<sup>3</sup>), PM<sub>2.5</sub> (15 µg/m<sup>3</sup>), or PM<sub>10</sub> (45 µg/m<sup>3</sup>). The 25th and 75th percentiles correspond to 121 and 222 days, respectively.
Abbreviations: SD, standard deviation; IQR, interquartile range.

### Exceedance days and cognitive trajectories

Based on models adjusted for pre-baseline age only, the estimated mean cognitive decline over 14 years in the analytic sample was -0.48 SD (95% CI, -0.51 to -0.46) for memory and - 0.24 SD (-0.27 to -0.20) for fluency.

In fully adjusted models, when considering all three pollutants together (Figure 2), there was no association between exceedance days and cognitive performance at baseline: the difference in baseline performance between the 75^th^ percentile and median of exceedance days was -0.03 SD (-0.07 to 0.01) for memory (p=0.20) and 0.01 SD (-0.04 to 0.06) for fluency (p=0.60). However, participants with more exceedance days had faster cognitive decline than those with fewer exceedance days. The difference in cognitive decline over 14 years between the 75^th^ percentile and the median number of exceedance days was -0.07 SD (-0.14 to 0.00) for memory (p=0.05) and -0.11 SD (-0.20 to -0.03) for fluency (p=0.007).

**Figure 2.**
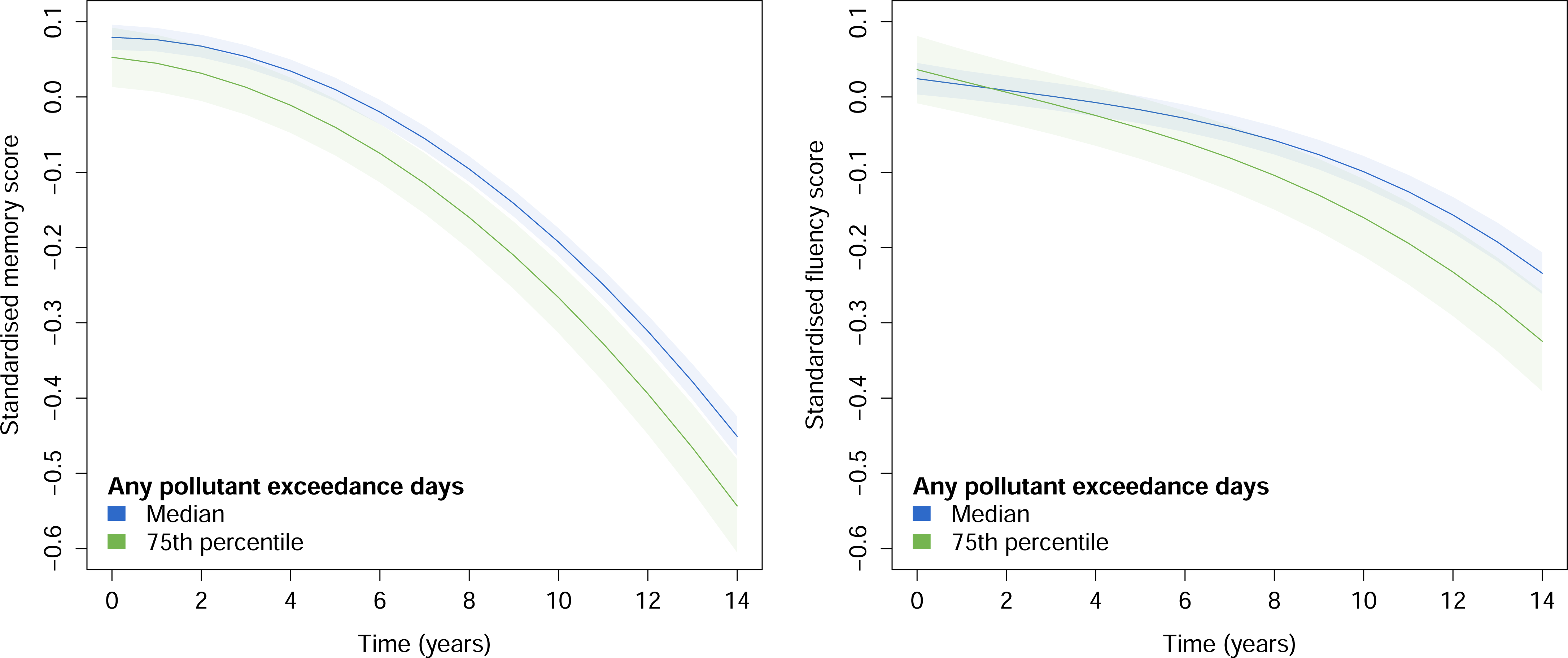
Predicted cognitive trajectories at median and 75^th^ percentile for exceedance days in any pollutant. Exceedance days refer to the number of days during the exposure year exceeding daily WHO guideline values for at least one of NO_2_ (25 g/m^3^), PM_2.5_ (15 g/m^3^), or PM_10_ (45 g/m^3^), with average cognitive trajectories estimated for the 50^th^ (170 days) and 75^th^ (222 days) percentiles. Estimates are from linear mixed models adjusted for age, birth year, sex, index of multiple deprivation, education level, wealth, urbanicity, and participant-specific mean annual NO_2_, PM_2.5_, and PM_10_ concentrations.

#### Pollutant-specific models

There was no association between NO_2_ exceedance days and cognitive performance at baseline in fully adjusted models (Figure 3). The difference in baseline memory performance between the 75^th^ percentile and median number of NO_2_ exceedance days was -0.01 SD (-0.06 to 0.03; p=0.57); the difference in baseline fluency performance was -0.01 SD (-0.06 to 0.05; p=0.83). However, more exceedance days for NO_2_ were associated with faster decline for both memory and fluency. The difference in cognitive decline over 14 years between the 75^th^ percentile and median of NO_2_ exceedance days was -0.09 SD (-0.17 to -0.01) for memory (p=0.02), and -0.10 SD (-0.19 to 0.00) for fluency (p=0.04).

**Figure 3.**
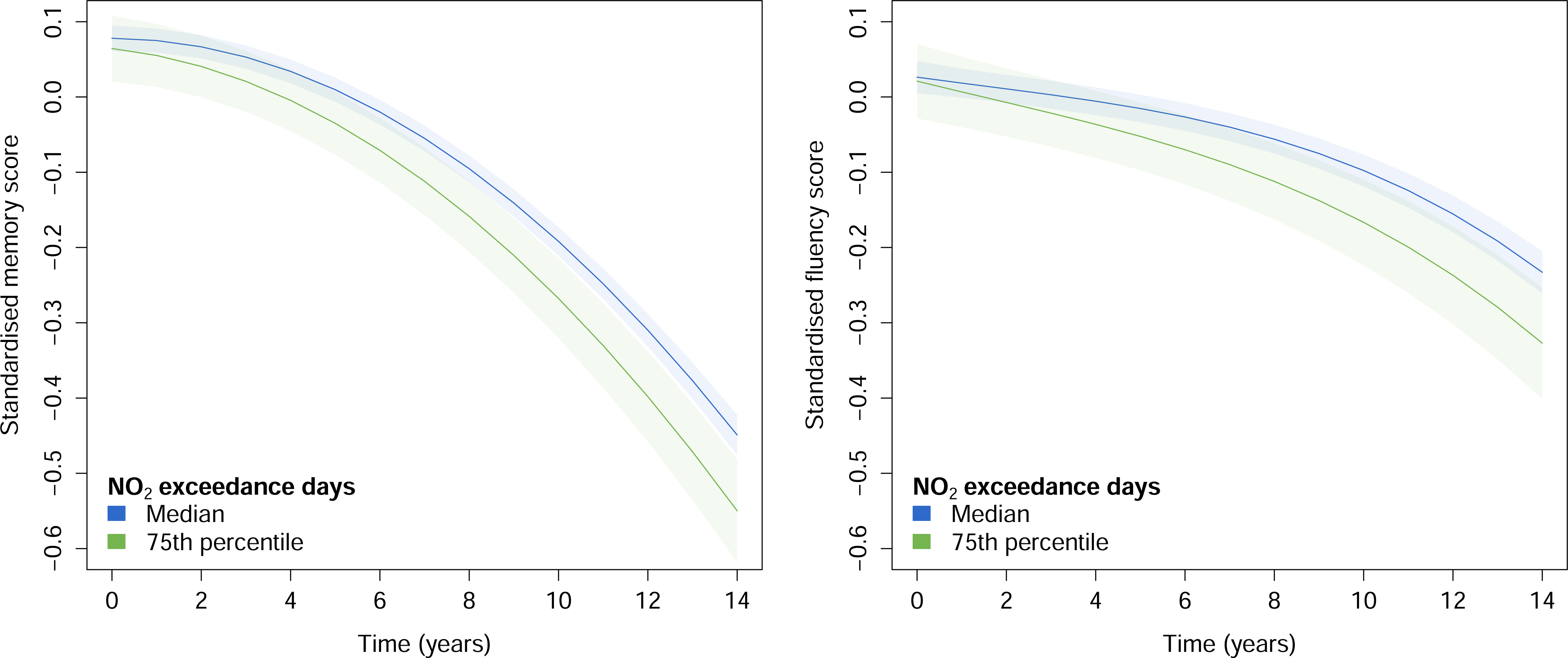
Predicted cognitive trajectories at median and 75^th^ percentile for NO_2_ exceedance days. Exceedance days refer to the number of days during the exposure year exceeding daily WHO guideline values for NO_2_ (25 µg/m^3^), with average cognitive trajectories estimated for the 50^th^ (149 days) and 75^th^ (212 days) percentiles. Estimates are from linear mixed models adjusted for age, birth year, sex, index of multiple deprivation, education level, wealth, urbanicity, and participant-specific mean annual NO_2_, PM_2.5_, and PM_10_ concentrations.

Neither PM_2.5_ exceedance days nor PM_10_ exceedance days were associated with baseline cognitive performance or subsequent cognitive decline in fully adjusted models (Figure 4). The difference in baseline memory performance between the 75^th^ percentile and median of exceedance days was 0.01 SD (-0.01 to 0.02) for PM_2.5_ (p=0.29) and 0.00 SD (-0.02 to 0.02) for PM_10_ (p=0.99). For fluency, the corresponding values were 0.01 SD (-0.01 to 0.02) for PM_2.5_ (p=0.38) and 0.01 SD (-0.01 to 0.02) for PM_10_ (p=0.49). The difference in memory decline over 14 years between the 75^th^ percentile and median of exceedance days was 0.01 SD (-0.01 to 0.02) for PM_2.5_ (p=0.56) and 0.02 SD (0.00 to 0.05) for PM_10_ (p=0.11). The corresponding difference in fluency decline was -0.01 SD (-0.03 to 0.01) for PM_2.5_ (p=0.53) and 0.01 SD (-0.01 to 0.04) for PM_10_ (p=0.31).

**Figure 4.**
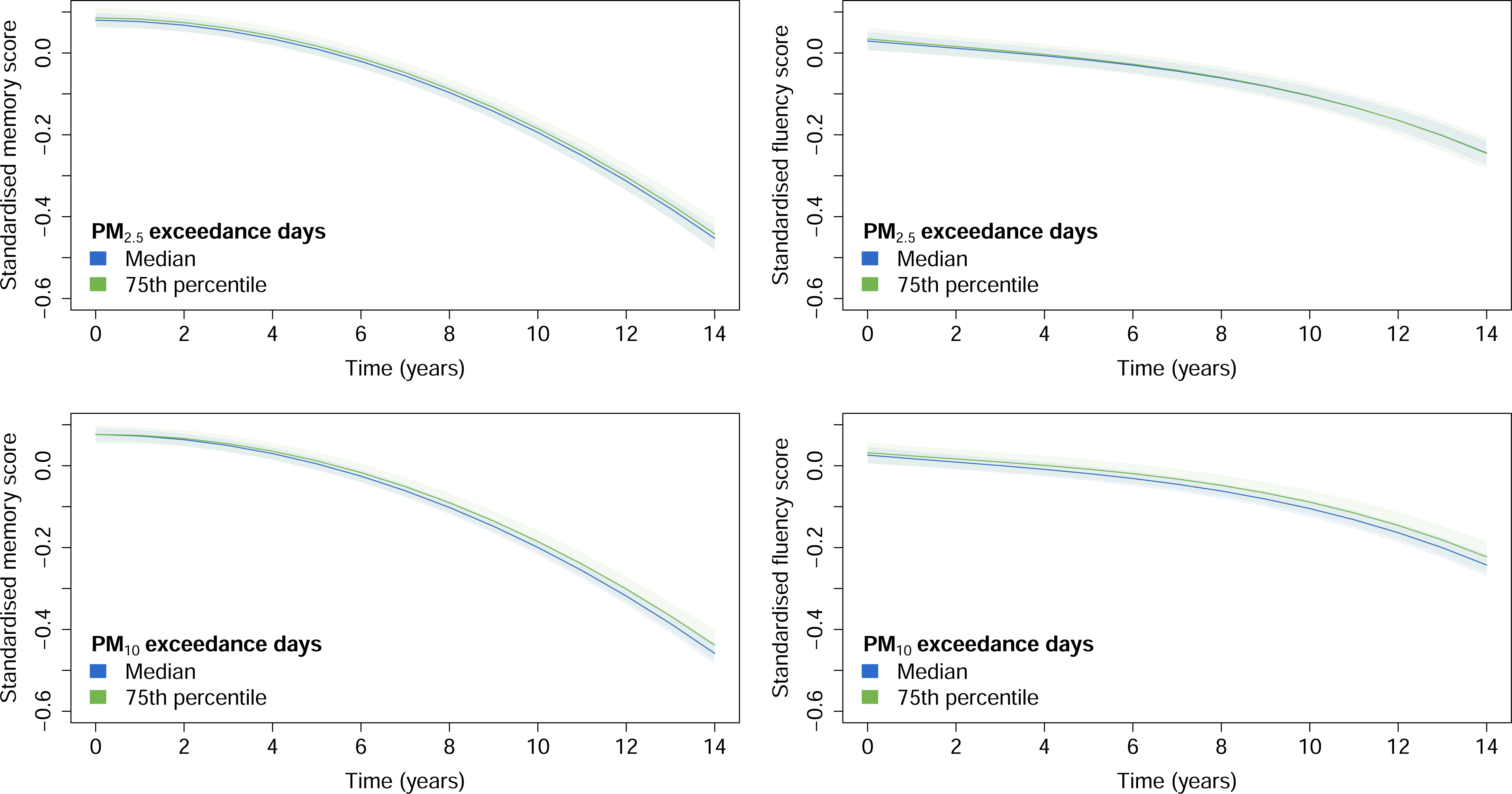
Predicted cognitive trajectories at median and 75^th^ percentile for particulate matter exceedance days. Exceedance days refer to the number of days during the exposure year exceeding daily WHO guideline values for particulate matter (15 µg/m^3^ for PM_2.5_, 45 µg/m^3^ for PM_10_), with average cognitive trajectories estimated for the 50^th^ (86 days for PM_2.5_, 21 days for PM_10_) and 75^th^ (94 days for PM_2.5_, 27 days for PM_10_) percentiles. Estimates are from linear mixed models adjusted for age, birth year, sex, index of multiple deprivation, education level, wealth, urbanicity, and participant-specific mean annual NO_2_, PM_2.5_, and PM_10_ concentrations.

### Additional analyses

There was no evidence that results for cognitive decline differed by sex (eTable 4), or baseline cognitive status (eTable 5). Results were unchanged when models were additionally adjusted for chronic conditions and health behaviors (N=12,166; eTable 6). Adjustment for time-varying air pollution exposure slightly strengthened NO_2_ associations with memory decline and attenuated associations with fluency, but point estimates remained very similar (eTable 7).

## Discussion

This study has three key findings. First, even in this low-to-moderate air pollution setting, recurrent daily exceedance of WHO guideline values was common for NO_2_, though somewhat less common for PM. Daily PM concentrations also varied less than NO_2_ concentrations, particularly for PM_2.5_. Second, after adjustment for mean annual exposure to air pollutants, participants exposed to more days during the year exceeding WHO daily guideline values for NO_2_ experienced faster cognitive decline. Finally, these associations were observed for NO_2_ but not PM_2.5_ or PM_10_. Together, these findings suggest that annual mean pollutant concentrations may obscure differences in day-to-day exposure patterning that are relevant to cognitive aging.

These results are in line with previous studies that suggest short-term peaks in air pollution are followed by poorer cognitive function.^5–7^ Our findings extend this evidence by showing that repeated high NO_2_ exposure across days is associated with longer-term cognitive decline even among individuals with similar annual mean pollutant exposure. However, no such associations were found for PM. One possible explanation for these results is that exceedance days capture different exposure processes for different pollutants. NO_2_ is strongly influenced by local combustion sources, particularly road traffic, heating, and power generation.^19^ These local high NO_2_ days could plausibly affect cognitive aging through acute vascular, inflammatory, or oxidative stress pathways, which have been proposed as mechanisms linking air pollution to neurodegeneration.^20^ PM_2.5_ reflects a more heterogeneous mixture of primary and secondary particles from local, regional, and transboundary sources: in many UK cities, agricultural emissions and long-range transport from continental Europe are the largest contributors to PM_2.5_.^21^ Consistent with this, daily PM_2.5_ concentrations varied substantially less than NO_2_ concentrations in our study, suggesting that PM_2.5_ exceedance days may more often reflect broader pollution episodes operating over larger spatial scales rather than repeated local high exposure days. In contrast to NO_2_ and PM_2.5_, the WHO daily guideline value for PM_10_ was high relative to observed concentrations in the analytic sample, resulting in a narrow exceedance distribution. PM_10_ exceedance days may therefore have been a less sensitive measure of relevant exposure patterning in this setting.

This study has several strengths. We used two decades of cognitive data from a large, nationally representative cohort linked to daily modelled air pollutant concentrations at participants’ residential locations. We used models that accounted for individual and geographic clustering, with detailed sociodemographic covariates. The study was undertaken in England, where air pollution concentrations are low to moderate by global standards, making the findings relevant to other European and North American settings where pollution levels are below those observed in many high exposure regions but still commonly exceed WHO daily guideline values. Finally, we defined high pollution days using exceedance of WHO daily guideline values, providing an interpretable measure of episodic exposure.

## Limitations

Requiring covariates and air pollution exposure to precede cognitive follow-up meant that the analytic baseline was generally later than participants’ first available cognitive assessment and participants with only one wave or without consecutive wave participation were excluded. Although this shortened follow-up, substantial follow-up remained because of the long duration of ELSA. Participants excluded from the analysis were somewhat more socioeconomically disadvantaged, and more generally, ELSA participants are 95% White, reflecting the demographic make-up of the target population,^22^ which may limit generalizability of findings to more disadvantaged or ethnically diverse populations. The missing-at-random assumption underpinning mixed models may not hold in this case as poorer cognitive function may drive attrition; however, follow-up duration was similar across levels of exceedance days, suggesting this issue is less likely to have strongly influenced the results. The exposure was measured during a single pre-baseline year, which may imperfectly represent participants’ longer-term pollution exposure patterns, particularly where exposure changed over time; residence-based estimates also do not capture exposure occurring away from home. Both may introduce exposure misclassification.

Analyses were limited to the cognitive domains assessed over a sufficiently long time period in ELSA to facilitate examination of long-term decline; associations with other cognitive domains should be examined. Finally, residual confounding remains possible despite adjustment for a broad range of individual- and area-level factors.

## Conclusions

In this cohort study of 12,235 middle-aged and older adults living in England, a higher number of NO_2_ exceedance days was associated with faster subsequent cognitive decline after adjustment for annual mean air pollutant concentrations. This suggests that daily exposure to high NO_2_ concentrations is related to accelerated cognitive aging even when offset by lower exposure on other days. Short-term high pollution episodes are already recognized as clinically relevant because they can trigger acute respiratory^23,24^ and cardiovascular events,^25–27^ with growing experimental evidence of negative impacts on short-term cognitive performance.^5,7^ Our findings suggest that recurrent high pollution exposure may also be important for longer-term cognitive health, even in settings with relatively low-to-moderate air pollution levels. Given the pollutant-specific nature of our findings, this study supports continued attention to local sources contributing to recurrent high NO_2_ exposure and policies such as low emission zones that target NO_2_ emissions.

## Data Availability

Data from the English Longitudinal Study of Ageing are available to researchers after registration with the UK data service at https://datacatalogue.ukdataservice.ac.uk/series/series/200011#abstract.

## Declaration of interests

The authors declare no competing interests.

## Funding/Support

The English Longitudinal Study of Ageing is funded by the National Institute on Aging (Ref: R01AG017644) and by a consortium of UK government departments: Department of Health and Social Care; Department for Transport; Department for Work and Pensions, which is coordinated by the National Institute for Health and Care Research (NIHR, Ref: 198-1074). The generation and linkage of air pollution data was carried out within the CONNECT Project (Wellcome Trust 320878/Z/24/Z).

## Role of the Funder/Sponsor

The funders had no role in the design and conduct of the study; collection, management, analysis, and interpretation of the data; preparation, review, or approval of the manuscript; and decision to submit the manuscript for publication.

## Access to Data and Data Analysis

Mikaela Bloomberg had full access to all the data in the study and takes responsibility for the integrity of the data and the accuracy of the data analysis.

## Notes

### Competing Interest Statement

The authors have declared no competing interest.

### Author Declarations

The English Longitudinal Study of Ageing received ethics approval from the South Central - Berkshire Research Ethics Committee. No further ethical approval is required for this secondary analysis.

### Summary of Updates

Corrected spelling of co-author name.

