## Supplementary materials for "Daily Exceedance of World Health Organization Air Pollutant Guidelines and Cognitive Aging"

**Supplemental materials**

### eMethods 1. Cognitive assessment.

The cognitive domains examined were episodic memory and verbal fluency, which show decline with dementia and are important for daily function.^1^ Memory was assessed using immediate and delayed recall tasks,^2^ in which participants were read a 10-word list and asked to recall it immediately (range: 0-10) and after a short delay (range: 0-10). These scores were summed to give a summary recall score (range: 0-20). Fluency was assessed using the animal naming task,^3^ where participants were asked to name out loud as many animals as possible in one minute. Memory was assessed at every wave and fluency, at all waves except wave 6. Cognitive scores were standardized using the distribution at the analytic baseline.

### eMethods 2. Index of multiple deprivation.

The Index of Multiple Deprivation (IMD) 2004 is a UK government produced area-level composite measure of relative deprivation, constructed from multiple domains that can be quantified for small geographic areas.^4^ It incorporates seven domains: income deprivation; employment deprivation; health deprivation; education, skills and training deprivation; barriers to housing and services; living environment deprivation and crime.

### eMethods 3. Model specification process.

We first determined the functional form of cognitive change using preliminary models adjusted for pre-baseline age and including polynomial terms for time. The appropriate degree of the time polynomial was determined through visual inspection of model-predicted cognitive trajectories together with Wald tests of the higher-order time terms. This supported linear and quadratic terms for time in the memory models and linear, quadratic, and cubic terms in the verbal fluency models.

We then fitted an a priori specified core model including exceedance days and all prespecified covariates (participant-specific annual mean NO_2_, PM_2.5_, and PM_10_ concentrations during the exposure year; pre-baseline age, birth year, sex, education, wealth quintile, IMD quintile, urbanicity), and interactions of the exposure and covariates with linear time.

Because the cognitive trajectories included higher-order time terms, we subsequently assessed whether corresponding higher-order interactions with the exposure or covariates were required. Including all possible quadratic and cubic time interactions would substantially increase the number of model parameters and therefore potentially reduce precision of estimates, so these terms were retained only where statistically significant by Wald tests (p<0.05). When a higher-order interaction was retained, all corresponding lower-order interaction terms were also retained. These tests were therefore used to determine the necessary complexity of the longitudinal trajectories and did not determine inclusion of the prespecified exposure or confounders.

Non-linearity in associations with exceedance days was assessed by adding quadratic exposure terms (exceedance days^2^, exceedance days^2^ x time). These coefficients were close to zero and not statistically significant by Wald tests in all models and were therefore not retained for model parsimony.

### eMethods 4. Sensitivity analyses.

To determine whether results differed by sex, we additionally included interactions between sex, exceedance days, and time in the models (sex x exceedance days, sex x exceedance days x time).

In line with previous work,^5^ participants were considered to show evidence of baseline cognitive impairment if they scored >1.5 standard deviations below the mean cognitive performance for their age (5-year bands) and education group (less than secondary, secondary, or above secondary). To determine whether results differed by cognitive status at baseline, we included cognitive status and interactions between cognitive status, exceedance days, and time (cognitive status, cognitive status x exceedance days, cognitive status x time, cognitive status x exceedance days x time).

We repeated analyses adjusted for self-reported physician-diagnosed chronic conditions (high blood pressure, diabetes, cancer, lung disease, heart disease, and stroke), physical activity (weekly moderate-to-vigorous physical activity or not), and smoking status (current smoker or not) reported during the pre-baseline wave.

To assess whether pollution exposure during cognitive follow-up influenced the observed associations, we repeated analyses additionally adjusting for individual-specific wave-specific mean NO_2_, PM_2.5_, and PM_10_ concentrations as time-varying covariates.

### eTable 1. Model specifications (fixed effects).

|  | ***Model terms*** | | | |
| --- | --- | --- | --- | --- |
|  | **Exposures** | **Core covariates** | **Additional covariates** | |
| ***Any pollutant*** | Exceedance days, exceedance days x time | time, age, age x time, birth year, birth year x time, sex, sex x time, education, education x time, wealth, wealth x time, IMD, IMD x time, urbanicity, urbanicity x time, mean NO_2_, mean NO_2_ x time, mean PM_2.5_, mean PM_2.5_ x time, mean PM_10_, mean PM_10_ x time | *Memory* | time^2^, age, age x time^2^, birth year x time^2^ |
|  |  |  | *Fluency* | time^2^, time^3^, age x time^2^, IMD x time^2^ |
| ***NO_2_*** | NO_2_ exceedance days, NO_2_ exceedance days x time |  | *Memory* | time^2^, age, age x time^2^, birth year x time^2^ |
|  |  |  | *Fluency* | time^2^, time^3^, age x time^2^, IMD x time^2^ |
| ***PM_2.5_*** | PM_2.5_ exceedance days, PM_2.5_ exceedance days x time |  | *Memory* | time^2^, age, age x time^2^, birth year x time^2^ |
|  |  |  | *Fluency* | time^2^, time^3^, age x time^2^, IMD x time^2^ |
| ***PM_10_*** | PM_10_ exceedance days, PM_10_ exceedance days x time |  | *Memory* | time^2^, age, age x time^2^, birth year x time^2^ |
|  |  |  | *Fluency* | time^2^, time^3^, age x time^2^, IMD x time^2^ |

Mean NO_2_, PM_2.5_, and PM_10_ refer to participant-specific annual mean during the exposure year.

### eTable 2. Participant characteristics in participants excluded versus included in the analytic sample.

|  | **In analytic sample** | | |  | | | **P-value** |
| --- | --- | --- | --- | --- | --- | --- | --- |
|  | *No*  (N=4,595) | *Yes*  (N=12,235) | |  | | |  |
| Age in years, mean (SD) | 61.7 (11.4) | | 62.5 (9.6) | |  | <0.001 | |
| Birth year, median (IQR) | 1947 (1933-1960) | | 1943 (1934-1950) | |  | <0.001 | |
| Sex |  | |  | |  |  | |
| Male | 2092 (46.0) | | 5541 (45.3) | |  |  | |
| Female | 2453 (54.0) | | 6694 (54.7) | |  | 0.40 | |
| Education level |  | |  | |  |  | |
| Less than secondary | 1931 (42.8) | | 5079 (41.5) | |  |  | |
| Secondary | 1898 (42.1) | | 5308 (43.4) | |  | 0.27 | |
| Above secondary | 678 (15.0) | | 1848 (15.1) | |  |  | |
| Wealth quintile |  | |  | |  |  | |
| 1 (least wealth) | 1261 (30.2) | | 2377 (19.4) | |  |  | |
| 2 | 912 (21.9) | | 2649 (21.7) | |  | <0.001 | |
| 3 | 717 (17.2) | | 2398 (19.6) | |  |  | |
| 4 | 618 (14.8) | | 2439 (19.9) | |  |  | |
| 5 (most wealth) | 664 (15.9) | | 2372 (19.4) | |  |  | |
| Neighbourhood deprivation |  | |  | |  |  | |
| 1 (least deprived) | 641 (17.3) | | 2382 (19.5) | |  |  | |
| 2 | 647 (17.5) | | 2322 (19.0) | |  | <0.001 | |
| 3 | 674 (18.2) | | 2366 (19.3) | |  |  | |
| 4 | 759 (20.5) | | 2502 (20.4) | |  |  | |
| 5 (most deprived) | 975 (26.4) | | 2663 (21.8) | |  |  | |
| Urbanicity |  | |  | |  |  | |
| Urban | 2846 (77.0) | | 9118 (74.5) | |  |  | |
| Town and fringe | 383 (10.4) | | 1450 (11.9) | |  | 0.007 | |
| Village/hamlet/isolated dwelling | 467 (12.6) | | 1667 (13.6) | |  |  | |
| Participant-specific annual mean air pollution, mean (SD) |  | |  | |  |  | |
| NO_2_ ($\mu$g/m^3^) | 24.7 (9.0) | | 25.2 (8.2) | |  | 0.004 | |
| PM_2.5_ ($\mu$g/m^3^) | 11.6 (2.0) | | 11.7 (1.6) | |  | <0.001 | |
| PM_10_ ($\mu$g/m^3^) | 21.8 (4.6) | | 22.6 (3.2) | |  | <0.001 | |
| Exceedance days, median (IQR) |  | |  | |  |  | |
| NO_2_ ($\mu$g/m^3^) | 137 (76-212) | | 149 (90-212) | |  | 0.001 | |
| PM_2.5_ ($\mu$g/m^3^) | 82 (45-93) | | 86 (68-94) | |  | <0.001 | |
| PM_10_ ($\mu$g/m^3^) | 18 (5-26) | | 21 (8-27) | |  | <0.001 | |

N (%) shown unless otherwise indicated. Denominators vary due to missing data.
Exceedance days refer to the number of days during the exposure year exceeding WHO guideline values for NO_2_ (25 $\mu$g/m^3^), PM_2.5_ (15 $\mu$g/m^3^), or PM_10_ (45 $\mu$g/m^3^). The 25th and 75th percentiles correspond to 121 and 222 days, respectively. Abbreviations: SD, standard deviation; IQR, interquartile range.

### eTable 3. Correlation across pollutants in participant-specific annual mean concentrations during the exposure year.

|  | ***NO_2_*** | ***PM_2.5_*** | ***PM_10_*** |
| --- | --- | --- | --- |
| ***NO_2_*** | 1.00 |  |  |
| ***PM_2.5_*** | 0.64 | 1.00 |  |
| ***PM_10_*** | 0.69 | 0.76 | 1.00 |

### eTable 4. Sex interaction terms.

| **Cognitive domain** | **Air pollutant** |  | **Sex x exceedance**  **days** | | |  | | **Sex x exceedance**  **days x time** | | |
| --- | --- | --- | --- | --- | --- | --- | --- | --- | --- | --- |
|  |  |  | *Coefficient*  *(95% CI)* | *P-value* |  | | *Coefficient*  *(95% CI)* | | *P-value* | |
| Memory | NO_2_ |  | 0.01 (-0.01 to 0.02) | 0.59 |  | | -0.01 (-0.04 to 0.02) | | | 0.52 |
|  | PM_2.5_ |  | 0.00 (0.00 to 0.01) | 0.29 |  | | 0.00 (-0.02 to 0.01) | | | 0.65 |
|  | PM_10_ |  | 0.01 (-0.01 to 0.02) | 0.36 |  | | 0.00 (-0.02 to 0.02) | | | 0.92 |
| Fluency | NO_2_ |  | 0.01 (-0.01 to 0.03) | 0.39 |  | | 0.00 (-0.03 to 0.03) | | | 0.87 |
|  | PM_2.5_ |  | 0.00 (0.00 to 0.01) | 0.32 |  | | 0.00 (-0.01 to 0.01) | | | 0.98 |
|  | PM_10_ |  | 0.01 (-0.01 to 0.02) | 0.35 |  | | 0.00 (-0.02 to 0.02) | | | 0.91 |

Exceedance days refer to the number of days exceeding WHO guideline values for air pollutants (25 $\mu$g/m^3^ for NO_2_, 15 $\mu$g/m^3^ for PM_2.5_, 45 $\mu$g/m^3^ for PM_10_). For consistency with the main models, sex x exceedance days coefficients are per increase in exceedance days from the 50^th^ to 75^th^ percentile (63 days for NO_2_, 8 days for PM_2.5_, and 6 days for PM_10_); sex x exceedance days x time coefficients are per 14 years. Estimates are from linear mixed models adjusted for age, birth year, sex, index of multiple deprivation, education level, wealth, urbanicity, and participat-specific annual mean NO_2_, PM_2.5_, and PM_10_ concentrations.

### eTable 5. Cognitive impairment interaction terms.

| **Cognitive domain** | **Air pollutant** |  | **Cognitive impairment x exceedance days** | | | |  | | | **Cognitive impairment x exceedance days x time** | | | |
| --- | --- | --- | --- | --- | --- | --- | --- | --- | --- | --- | --- | --- | --- |
|  |  |  | *Coefficient*  *(95% CI)* | *P-value* | |  | | | *Coefficient*  *(95% CI)* | | | *P-value* | |
| Memory | NO_2_ |  | -0.01 (-0.03 to 0.00) | | 0.15 | | |  | | | 0.00 (-0.04 to 0.03) | | 0.80 |
|  | PM_2.5_ |  | 0.01 (0.00 to 0.01) | | 0.16 | | |  | | | -0.01 (-0.02 to 0.01) | | 0.24 |
|  | PM_10_ |  | 0.01 (0.00 to 0.02) | | 0.14 | | |  | | | -0.01 (-0.03 to 0.01) | | 0.35 |
| Fluency | NO_2_ |  | -0.03 (-0.05 to 0.00) | | 0.02 | | |  | | | -0.02 (-0.05 to 0.02) | | 0.31 |
|  | PM_2.5_ |  | 0.00 (-0.01 to 0.01) | | 0.71 | | |  | | | 0.00 (-0.02 to 0.01) | | 0.55 |
|  | PM_10_ |  | 0.01 (0.00 to 0.03) | | 0.05 | | |  | | | -0.01 (-0.04 to 0.01) | | 0.24 |

Exceedance days refer to the number of days exceeding WHO guideline values for air pollutants (25 $\mu$g/m^3^ for NO_2_, 15 $\mu$g/m^3^ for PM_2.5_, 45 $\mu$g/m^3^ for PM_10_). For consistency with the main models, cognitive impairment x exceedance days coefficients are per increase in exceedance days from the 50^th^ to 75^th^ percentile (63 days for NO_2_, 8 days for PM_2.5_, and 6 days for PM_10_); cognitive impairment x exceedance days x time coefficients are per 14 years. Estimates are from linear mixed models adjusted for age, birth year, sex, index of multiple deprivation, education level, wealth, urbanicity, and participant-specific annual mean NO_2_, PM_2.5_, and PM_10_ concentrations.

### eTable 6. Association of exceedance days with baseline cognitive performance and 14-year cognitive decline with additional adjustment for health-related variables (N=12,166).

| **Cognitive domain** | **Air pollutant** |  | **Baseline cognitive performance** | | |  | | **14-year cognitive decline** | |
| --- | --- | --- | --- | --- | --- | --- | --- | --- | --- |
|  |  |  | *Coefficient*  *(95% CI)* | *P-value* |  | | *Coefficient*  *(95% CI)* | | *P-value* |
| Memory | NO_2_ |  | 0.00 (-0.01 to 0.02) | 0.67 |  | | -0.09 (-0.16 to -0.01) | | 0.03 |
|  | PM_2.5_ |  | -0.02 (-0.06 to 0.03) | 0.53 |  | | 0.00 (-0.01 to 0.02) | | 0.62 |
|  | PM_10_ |  | 0.01 (0.00 to 0.02) | 0.27 |  | | 0.02 (0.00 to 0.05) | | 0.08 |
| Fluency | NO_2_ |  | 0.00 (-0.02 to 0.01) | 0.75 |  | | -0.09 (-0.18 to 0.00) | | 0.04 |
|  | PM_2.5_ |  | -0.01 (-0.06 to 0.05) | 0.83 |  | | -0.01 (-0.03 to 0.01) | | 0.61 |
|  | PM_10_ |  | 0.01 (-0.01 to 0.02) | 0.41 |  | | 0.02 (-0.01 to 0.04) | | 0.29 |

Exceedance days refer to the number of days exceeding WHO guideline values for air pollutants (25 $\mu$g/m^3^ for NO_2_, 15 $\mu$g/m^3^ for PM_2.5_, 45 $\mu$g/m^3^ for PM_10_). Estimates compare the 75^th^ (212 days for NO_2_, 94 days for PM_2.5_, 27 days for PM_10_) with the 50^th^ (149 days for NO_2_, 86 days for PM_2.5_, 21 days for PM_10_) percentiles for exceedance days. Based on linear mixed models adjusted for age, birth year, sex, index of multiple deprivation, education level, wealth, urbanicity, and participant-specific annual mean NO_2_, PM_2.5_, and PM_10_ concentrations. Additionally adjusted for self-reported baseline chronic conditions, smoking status, and physical activity.

### eTable 7. Association of exceedance days with baseline cognitive performance and 14-year cognitive decline with additional adjustment for post-exposure air pollution concentrations.

| **Cognitive domain** | **Air pollutant** |  | **Baseline cognitive performance** | | |  | | **14-year cognitive decline** | |
| --- | --- | --- | --- | --- | --- | --- | --- | --- | --- |
|  |  |  | *Coefficient*  *(95% CI)* | *P-value* |  | | *Coefficient*  *(95% CI)* | | *P-value* |
| Memory | NO_2_ |  | 0.01 (-0.01 to 0.03) | 0.40 |  | | -0.10 (-0.17 to -0.02) | | 0.01 |
|  | PM_2.5_ |  | -0.01 (-0.06 to 0.04) | 0.67 |  | | 0.00 (-0.01 to 0.02) | | 0.62 |
|  | PM_10_ |  | 0.01 (-0.00 to 0.02) | 0.22 |  | | 0.02 (-0.01 to 0.04) | | 0.15 |
| Fluency | NO_2_ |  | 0.00 (-0.01 to 0.02) | 0.70 |  | | -0.08 (-0.17 to 0.01) | | 0.07 |
|  | PM_2.5_ |  | -0.01 (-0.06 to 0.05) | 0.79 |  | | -0.01 (-0.03 to 0.01) | | 0.42 |
|  | PM_10_ |  | 0.01 (-0.00 to 0.02) | 0.21 |  | | 0.01 (-0.02 to 0.04) | | 0.35 |

Exceedance days refer to the number of days exceeding WHO guideline values for air pollutants (25 $\mu$g/m^3^ for NO_2_, 15 $\mu$g/m^3^ for PM_2.5_, 45 $\mu$g/m^3^ for PM_10_). Estimates compare the 75^th^ (212 days for NO_2_, 94 days for PM_2.5_, 27 days for PM_10_) with the 50^th^ (149 days for NO_2_, 86 days for PM_2.5_, 21 days for PM_10_) percentiles for exceedance days. Based on linear mixed models adjusted for age, birth year, sex, index of multiple deprivation, education level, wealth, urbanicity, and participant-specific annual mean NO_2_, PM_2.5_, and PM_10_ concentrations. Additionally adjusted for individual-specific wave-specific mean NO_2_, PM_2.5_, and PM_10_ concentrations as time-varying covariates.

### eFigure 1. Illustrative timing of covariate, air pollution exposure, and cognitive outcome assessment.

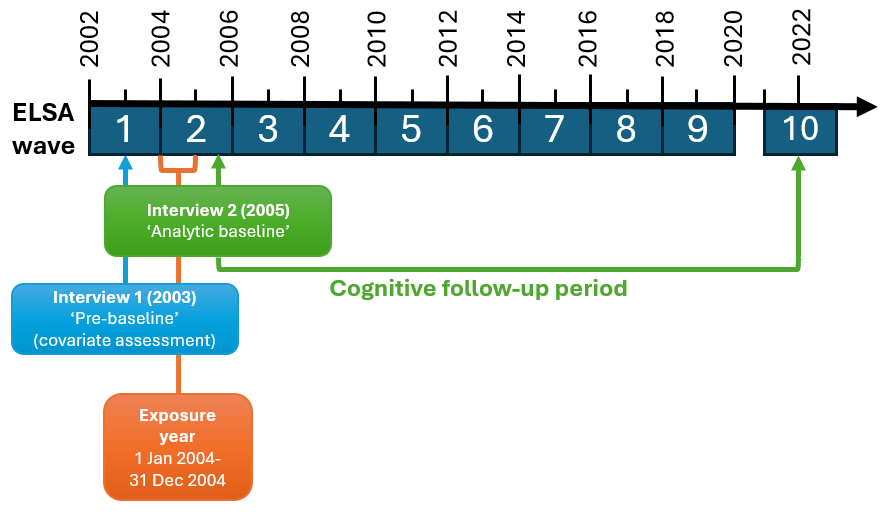

Example for a participant whose first cognitive assessment meeting the study eligibility criteria occurred at wave 2. The immediately preceding wave (wave 1) was defined as pre-baseline and used for covariate assessment. Air pollution exposure was assessed during the calendar year preceding analytic baseline (2004), and the wave 2 interview in 2005 was defined as analytic baseline. Cognitive follow-up began at analytic baseline and continued through subsequent waves.
